# Efficacy of Tapering Biologics and JAK Inhibitors in Rheumatoid Arthritis: A Systematic Review and Meta-analysis

**DOI:** 10.1101/2025.09.29.25336875

**Authors:** Sandra Rotea-Salvo, Laura Galindo-Domínguez, Belén Acasuso-Pardo de vera, Vanesa Balboa-Barreiro, Luis Ramudo-Cela, Maite Silva-Díaz, Natividad Oreiro-Villar, Francisco J. De Toro-Santos, Isabel Martín-Herranz, Francisco J. Blanco-García

## Abstract

This study assessed the evidence on the efficacy of tapering anti-TNF, JAK inhibitors, and tocilizumab in rheumatoid arthritis patients. In February 2024, a systematic review was conducted by searching Medline, Embase, Web of Sciences, and the Cochrane Library for randomized controlled trials comparing the efficacy of tapering vs standard treatment. The outcomes evaluated were the maintenance of low disease activity (LDA), remission, and flare-ups. A meta-analysis was conducted when data were available. The risk of bias was assessed using RoB 2. The study was registered on PROSPERO. A total of 2861 records were identified, with 1638 records screened after removing duplicates. Finally, fifteen studies involving 2782 patients were included. Follow-up ranged from 6 months to 3.5 years. Tapering anti-TNF did not affect LDA maintenance while showing a lower probability of maintaining remission (RR 0.69, 95% CI 0.57-0.84) and a higher risk of flare-ups (RR 1.96, 95% CI 1.57-2.45). Tapering JAK inhibitors showed a decreased probability of maintaining LDA (RR 0.83, 95% CI 0.76-0.91) and remission (RR 0.86, 95% CI 0.75-0.99), and more frequent and earlier flares. Tapering tocilizumab also resulted in a lower probability of maintaining LDA or remission and a higher risk of flares. Although tapering anti-TNF did not affect LDA maintenance, due to the increased risk of flare-ups and reduced remission probability, routine dose tapering of anti-TNF, JAK inhibitors, and tocilizumab for all patients is not recommended. Identifying patients who may benefit from tapering is crucial.

## Introduction

Rheumatoid arthritis (RA) is a chronic inflammatory autoimmune disease that primarily affects the joints, but it can also present extra-articular manifestations. Advances in disease assessment, therapeutic strategies, and new drugs have transformed the management of this disease, markedly enhancing the quality of life for patients [1].

The current approach is based on a treat-to-target strategy aiming to achieve remission or low disease activity (LDA) as soon as possible. Once the desired outcome has been reached, tapering disease-modifying antirheumatic drugs (DMARDs) may be feasible [2,3].

Various studies, including randomized controlled trials (RCTs) [4], observational studies [5], and real-world studies [6] have suggested that dose reduction could be successful in some patients. Some previous systematic reviews have shown it is feasible to reduce DMARDs without compromising the clinical response [7,8]. International [9,10] and national guidelines [11] provide recommendations about tapering. EULAR agrees that tapering DMARDs may be considered when the patient has achieved remission or LDA for at least 6 months and after discontinued glucocorticoids [9].

The ACR conditionally recommends the continuation of all DMARDs over a dose reduction, citing evidence that patients who reduce their DMARD have a higher risk of flare compared to those who maintain their original dose [10]. Additionally, some RCTs [12,13] indicate that tapering DMARDs is associated with an increased risk of flares and have not demonstrated non-inferiority for biological tapering. Overall, current guidelines emphasize that tapering decisions should be individualized, relying on clinical judgment and involving shared decision-making with patients.

Based on the contradictory data regarding tapering, this manuscript aims to update the analysis of tapering efficacy for anti-TNF agents in RA, incorporating new RCTs. Additionally, this meta-analysis and systematic review uniquely include studies on the tapering efficacy of JAK inhibitors and tocilizumab, which, to our knowledge, has not been done before. These drugs are among the most widely used in hospitals for adult RA patients when conventional synthetic DMARDs (csDMARDs) are ineffective or not tolerated. This comprehensive approach offers new insights into optimizing treatment strategies in RA management.

## Methods

### Search strategy

This systematic review and meta-analysis followed PRISMA guidelines [14,15] and was registered on PROSPERO (CRD42024509208).

We searched on February 8, 2024, Medline, Embase, Web of Sciences, and the Cochrane Library. References of relevant publications were manually searched to identify potential studies that might have been missed in the electronic search. No language or time restrictions were applied. Search terms used included “Rheumatoid arthritis”, “Anti-TNF”, “JAK inhibitors”, “Tocilizumab”, “Down titration”, “Reduction” and “Tapering”. The complete search strategy of each database is shown in the Supplementary Table S1.

### Eligibility criteria and study selection

Studies were required to compare an intervention-tapering group, which involved a dose-reduction strategy for the study drugs (anti-TNF, JAK inhibitors, and tocilizumab), with a control-no tapering group, which continued with the standard treatment. Tapering was defined as lowering the dose or spacing the interval of administration, including tapering to stop after a gradual reduction. The included studies had to meet the following criteria:

– *Population:* adult patients (≥18 years old) diagnosed with RA and treated with anti-TNF, JAK inhibitors, or tocilizumab who achieved remission or LDA.
– *Intervention*: a tapering strategy for the study drugs according to the definition mentioned above.
– *Comparator*: a standard-dose treatment as per labelled doses for RA. Studies with more than two arms (in addition to the control and intervention groups) were included, such as those with a study drug discontinuation arm (stopped group). However, only data from the control and tapering groups were recorded.
– *Outcomes*: efficacy of both strategies, analyzing LDA, remission, and flare-ups.
– *Design of studies:* RCTs.

Only original research articles were included. Other publication types such as letters, editorials, conference abstracts, or sub-analyses of original studies were excluded. Studies with a control group compared to an intervention group involving total cessation (stop) of study drugs without gradual discontinuation were also excluded.

Two authors (SRS, and LGD) independently reviewed and selected the included studies. In cases of discrepancies, a third author (BAP) was consulted.

### Data extraction

The following data were extracted from all included studies: author, study name, year of publication, population, study design, drug and strategy, clinical status before tapering, endpoint, main outcome, and results.

### Data synthesis and data analysis

A meta-analysis was conducted using the available data for effect measures (maintenance of LDA, remission, or flare-ups) and drug category (anti-TNF and JAK inhibitors). The meta-analysis was performed using a meta-R package and required a minimum of two studies. Risk ratios (RR) and 95% confidence intervals (CI) were calculated for each study. Heterogeneity was assessed using the I^2^ test. When a meta-analysis could not be performed, a narrative synthesis of the results was provided, comparing the standard group (SG) with the reduced group (RG).

### Study risk of bias assessment

The Cochrane risk-of-bias tool for RCT (RoB 2) was employed to assess the risk of bias (high, low, or some concerns) based on the randomization process, deviations from intended interventions, missing outcome data, measurement of the outcome and selection of the reported result [16]. Two researchers (SRS and LGD) independently assessed the risk of bias. Disagreements were resolved by a third researcher (BAP).

## Results

### Study Selection

A total of 2861 records were identified. After removing duplicates, 1638 records were screened based on title and abstract, and 75 were reviewed in full text. Finally, 15 studies [4,12,13,17-28] were included in the systematic review (Figure 1).

**Figure 1.**
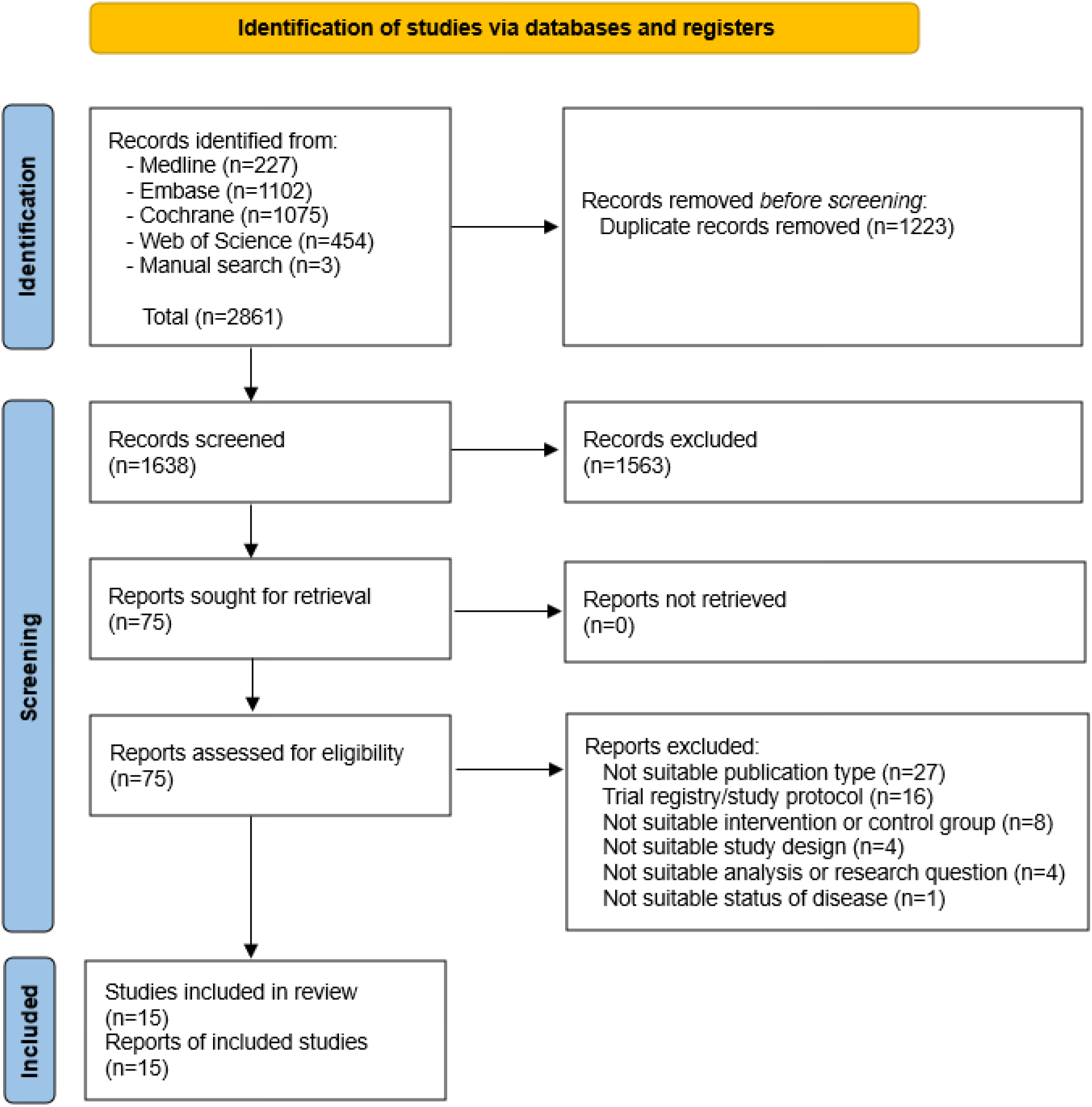
PRISMA flow diagram for identification of studies.

### Study characteristics

The characteristics of the included studies are summarized in Table 1. A total of 2782 participants were enrolled, predominantly female (60%-87%) with a mean age from 47 to 59 years. The study by El Miedany et al. [17] did not report sociodemographic details, and in the study by Kedra et al. [12], sociodemographic characteristics were not presented separately by treatment. The follow-up period varied between 6 months to 3.5 years.

**Table 1.**
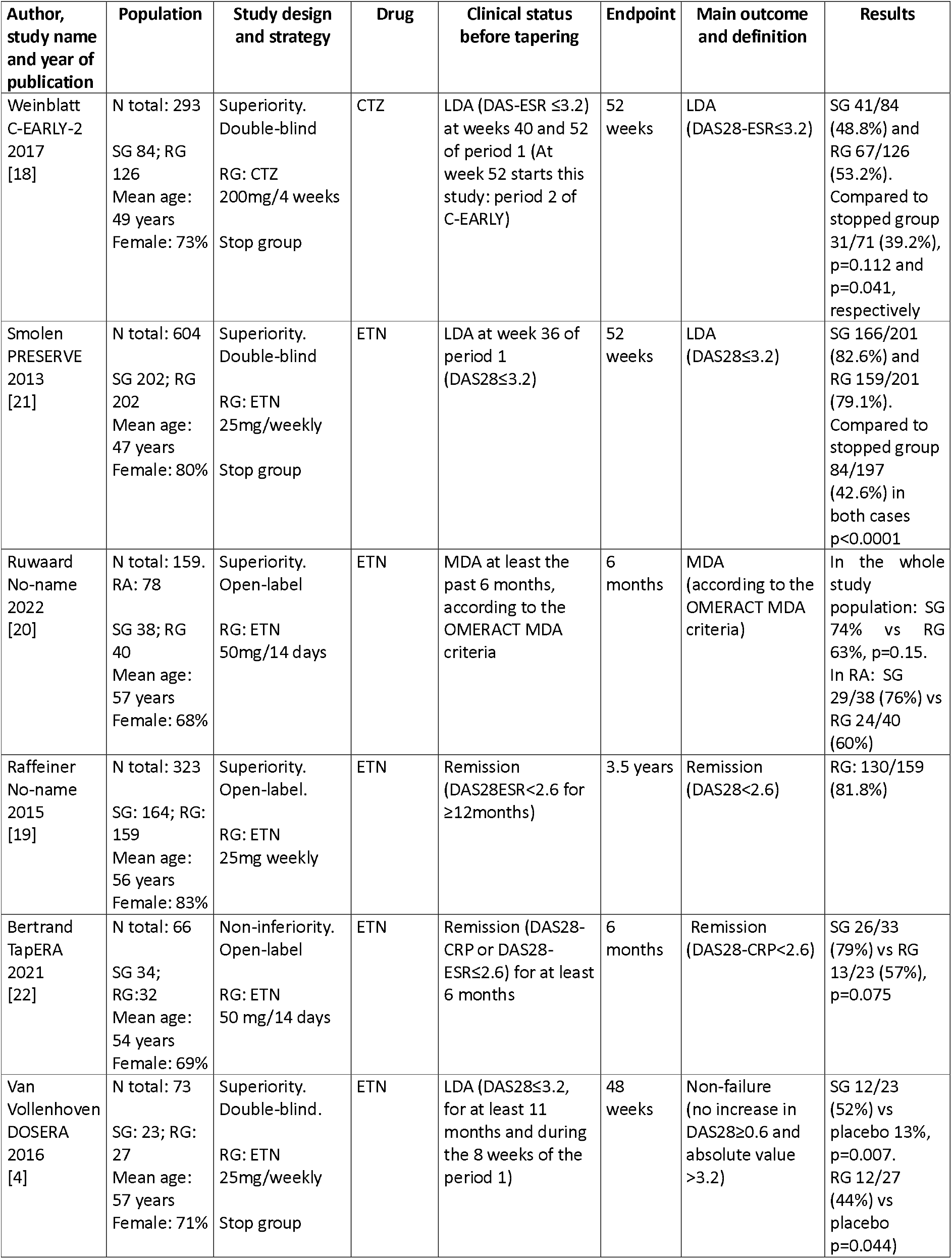
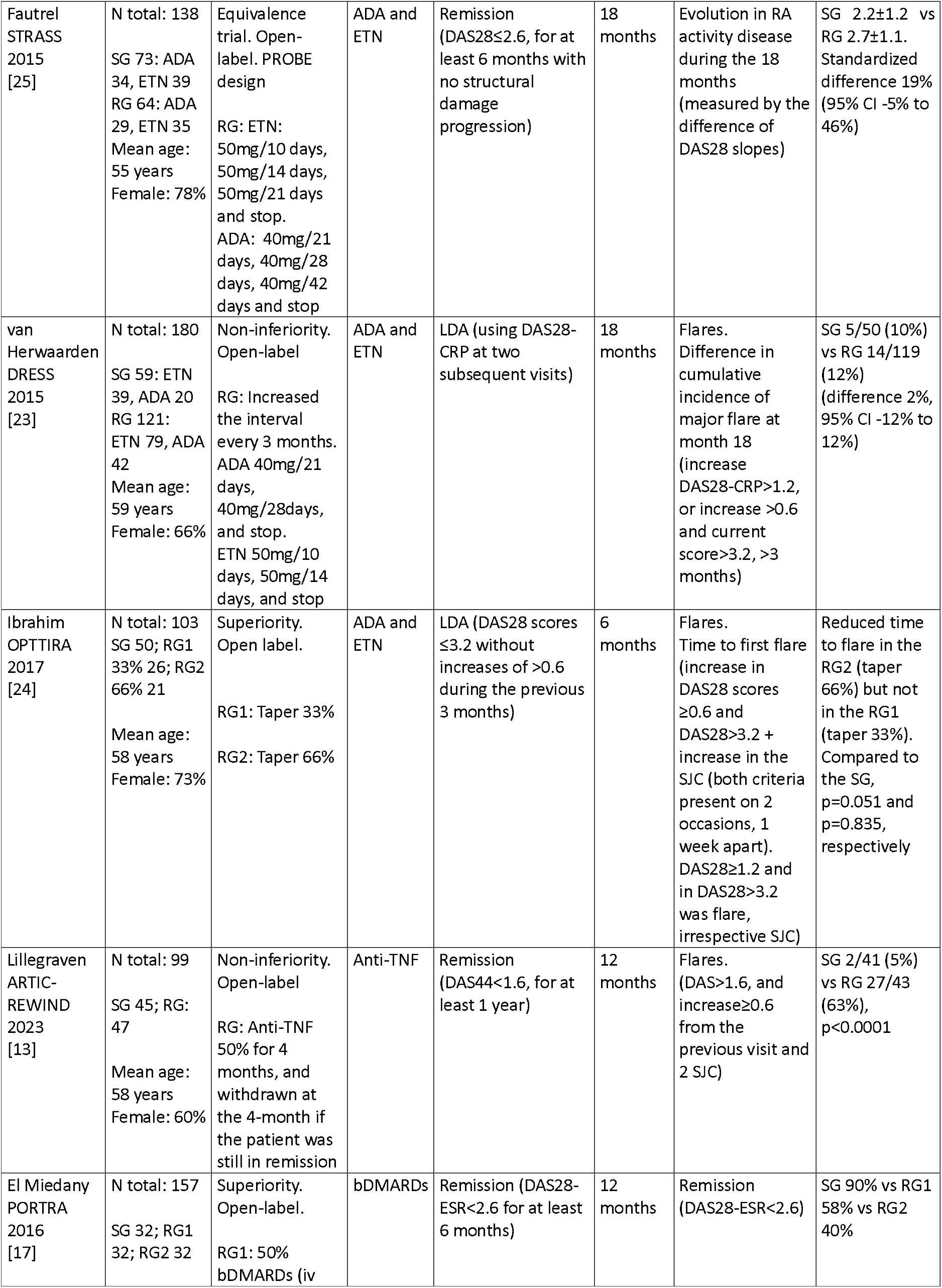
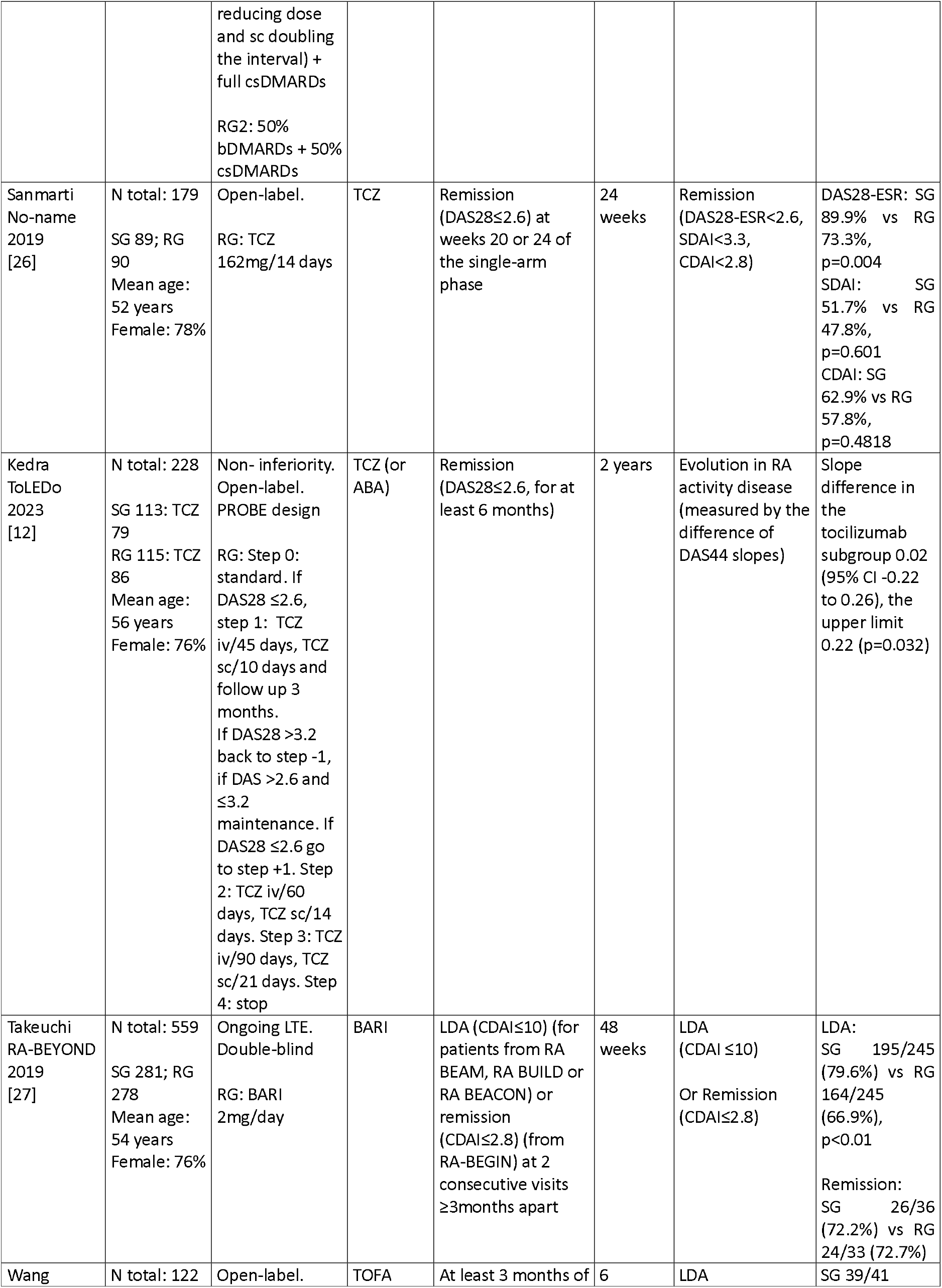
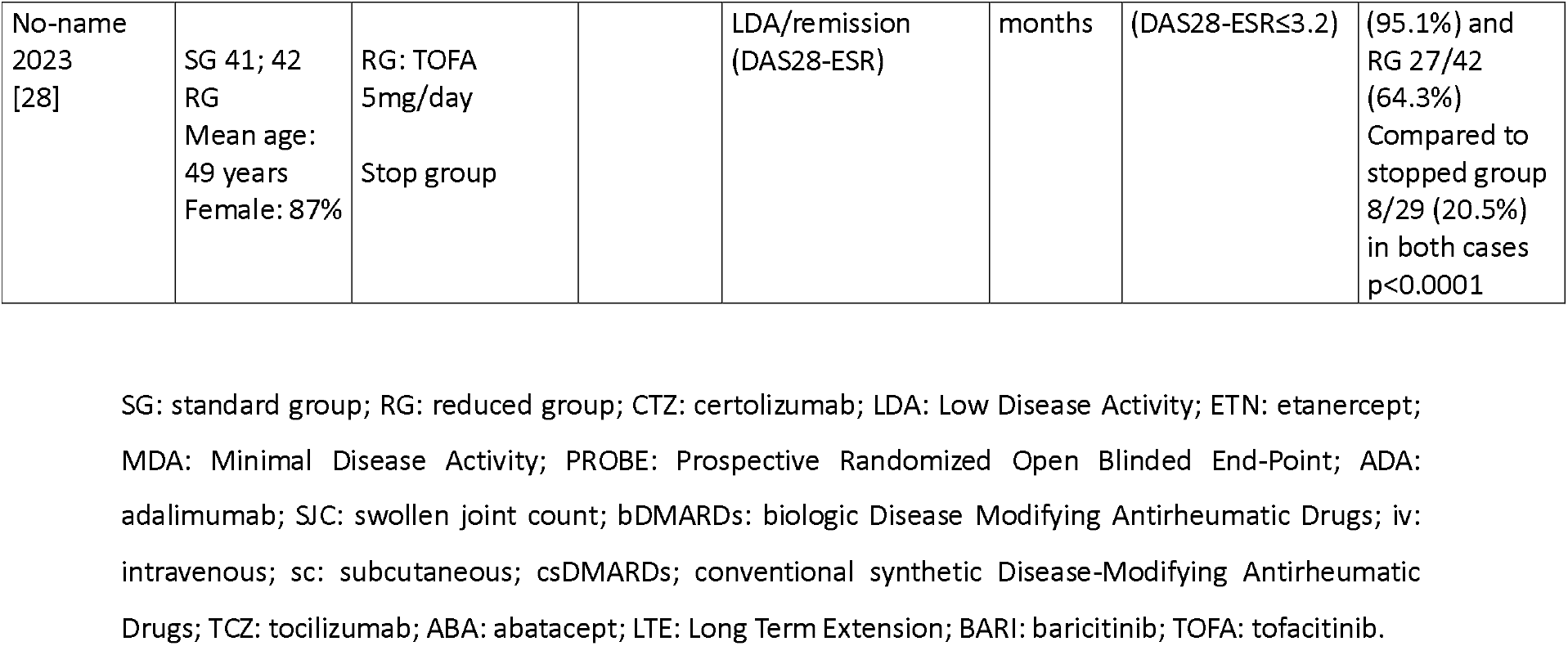
Characteristics of the included studies according to drug class and main outcome.

The evaluated drugs included certolizumab [18], etanercept [4, 19-22], adalimumab or etanercept [23,24,25], anti-TNF [13,17], tocilizumab [12,17,26], baricitinib [27], and tofacitinib [28]. The PORTRA study [17] included patients treated with anti-TNF and tocilizumab, and eight patients with abatacept, although it was not specified in which arm they were. Given the small number of abatacept-treated patients, we decided to include this study in our review.

Most studies compared two treatment arms (SG vs RG). The OPTTIRA study [24] employed three arms: SG and two RG (tapering by 33% and 66%), while PORTRA [17] reported five arms (groups 1, 2, and 5 included in this review, referring to taper biologic DMARDs (bDMARDs) and full csDMARDs, taper both bDMARDs and csDMARDs, and SG, respectively). Furthermore, four studies included a third arm exploring drug discontinuation alongside control and reduction arms [4,18,21,28].

Various optimization strategies were employed across the studies, including fixed-dose reduction [4,13,17-22,24,26-28], gradual dose reduction [12,23,25], and complete discontinuation attempted in four studies [12,13,23,25].

### Risk of bias in studies

Four studies presented a low risk of bias in all items, two had some concerns, and the other nine high risk. The main domain in which the studies presented a high risk of bias was the related measurement of the outcome, due to the open-label design (Figure 2).

**Figure 2.**
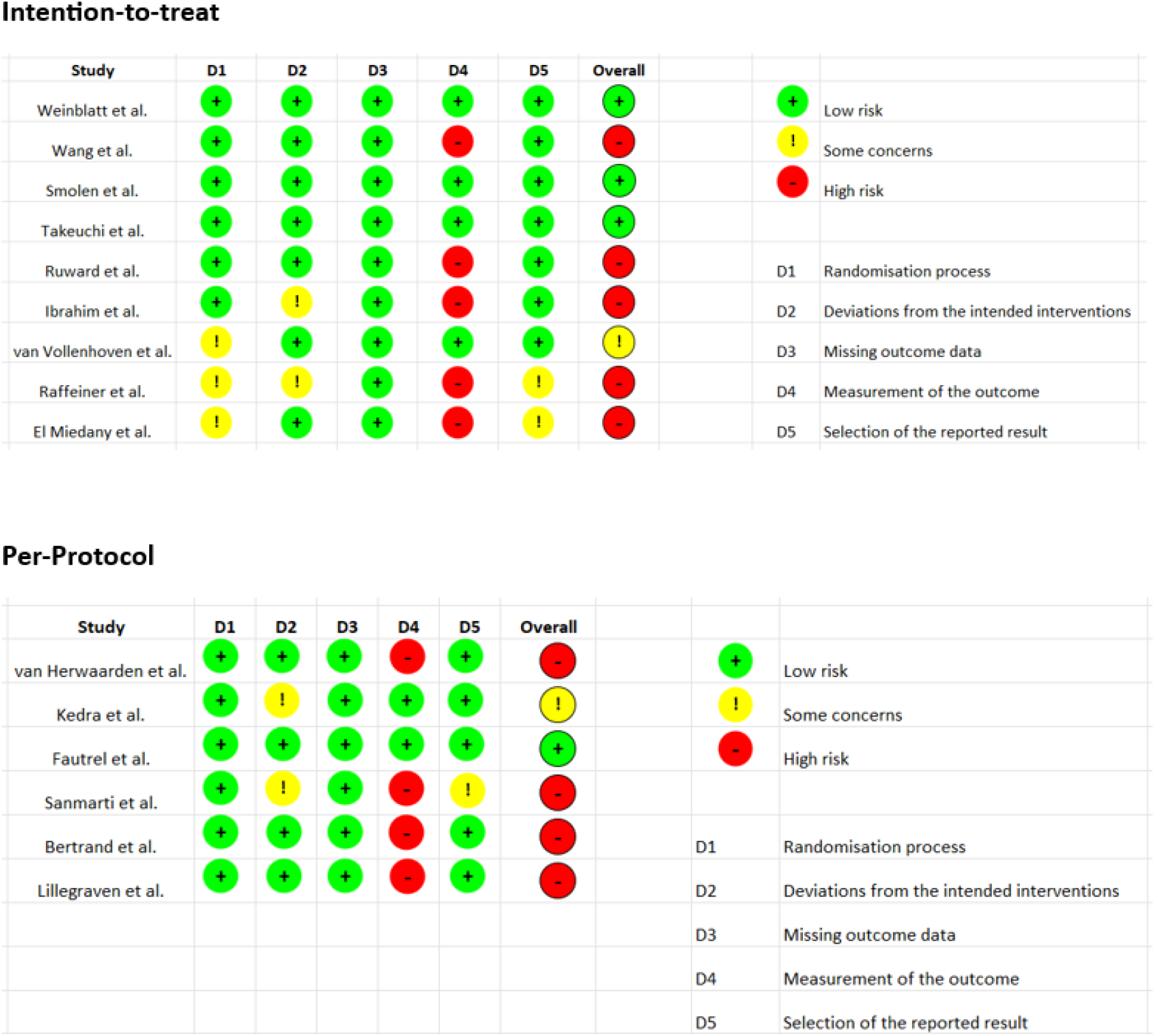
Risk of bias of the included studies.

### Outcomes of anti-TNF

1. **LDA:** a pooled analysis of four studies revealed that tapering anti-TNF did not affect the maintenance of LDA (RR 0.96, 95% CI 0.87-1.05) with no heterogeneity (I^2^ 0%) (Figure 3a).
2. **Remission:** three studies showed a statistically significant reduction in the probability of maintaining remission when tapering anti-TNF (RR 0.69, 95% CI 0.57-0.84) with high heterogeneity (I^2^ 57%) (Figure 3b).
3. **Flares-up:** seven studies revealed a statistically significant association between tapering anti-TNF and an increased risk of flare-ups (RR 1.96, 95% CI 1.57-2.45) with high heterogeneity (I^2^ 73%) (Figure 3c).

**Figure 3.**
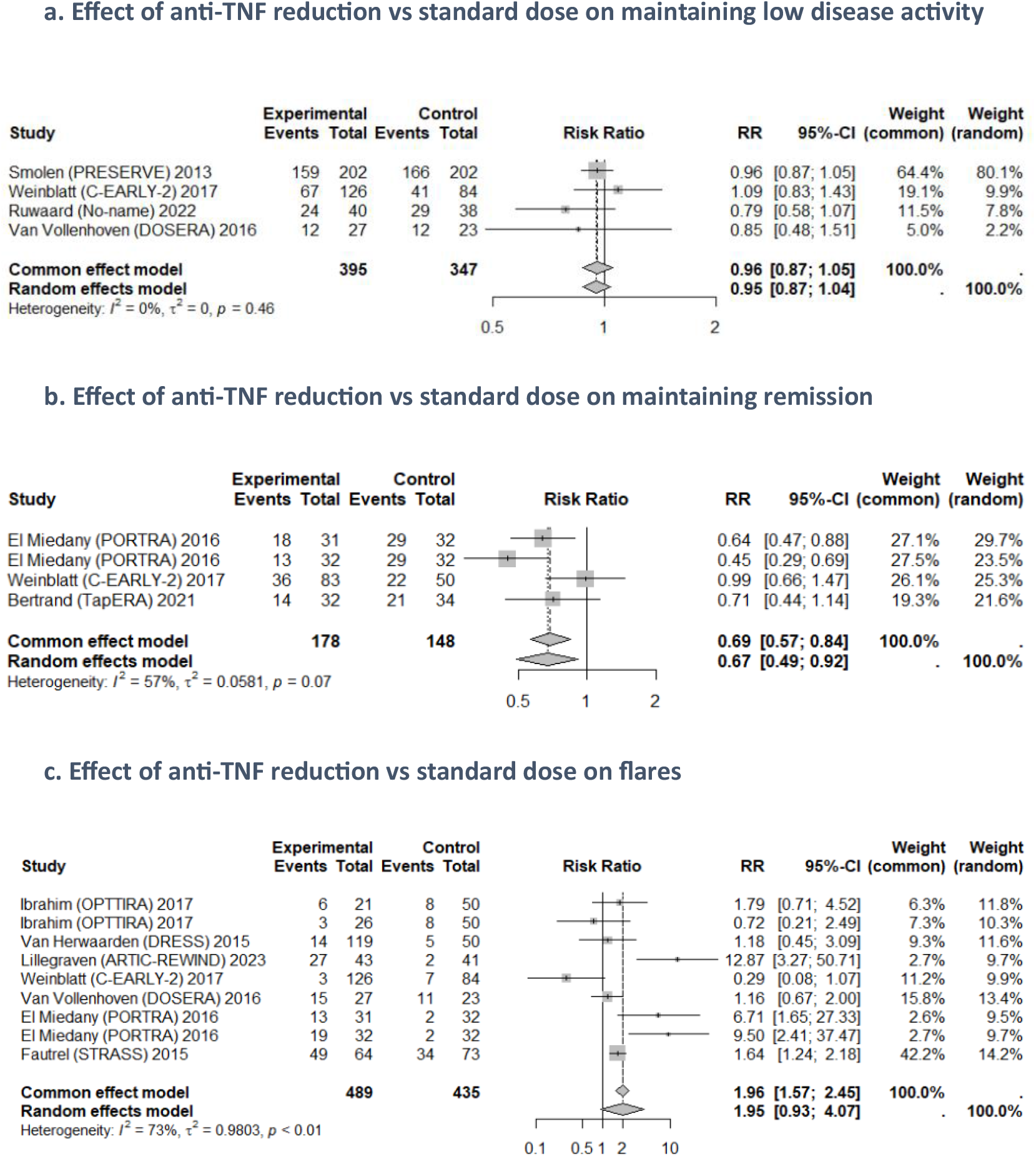
Effect on disease activity comparing anti-TNF tapering strategy to maintenance full-treatment. RR: Risk ratio; CI: Confidence interval.

### Outcomes of JAK inhibitors

1. **LDA:** two studies showed a reduced likelihood of maintaining LDA in the tapering group compared to the control group (RR 0.83, 95% CI 0.76-0.91) although with high heterogeneity (I^2^ 71%) (Figure 4a).
2. **Remission:** results from two studies also showed that tapering was associated with a reduced likelihood of maintaining remission (RR 0.86, 95% CI 0.75-0.99) with no heterogeneity (I^2^ 0%) (Figure 4b).
3. **Flares-up:** a meta-analysis was not performed because the data were not included in the paper. A systematic review of the manuscripts reported that Takeuchi et al. [27] observed earlier and more frequent relapses with baricitinib-reduced dose (SG 23% vs RG 37%, p=0.001). Wang et al. [28] reported a higher proportion of flares in the tofacitinib-reduced group [4/41 (9.8%) vs 15/42 (35.7%)], a modest loss of efficacy (log-rank, p=0.0039), and a shorter mean time to flare in the RG compared to SG (5.8 months vs 4.7 months).

**Figure 4.**
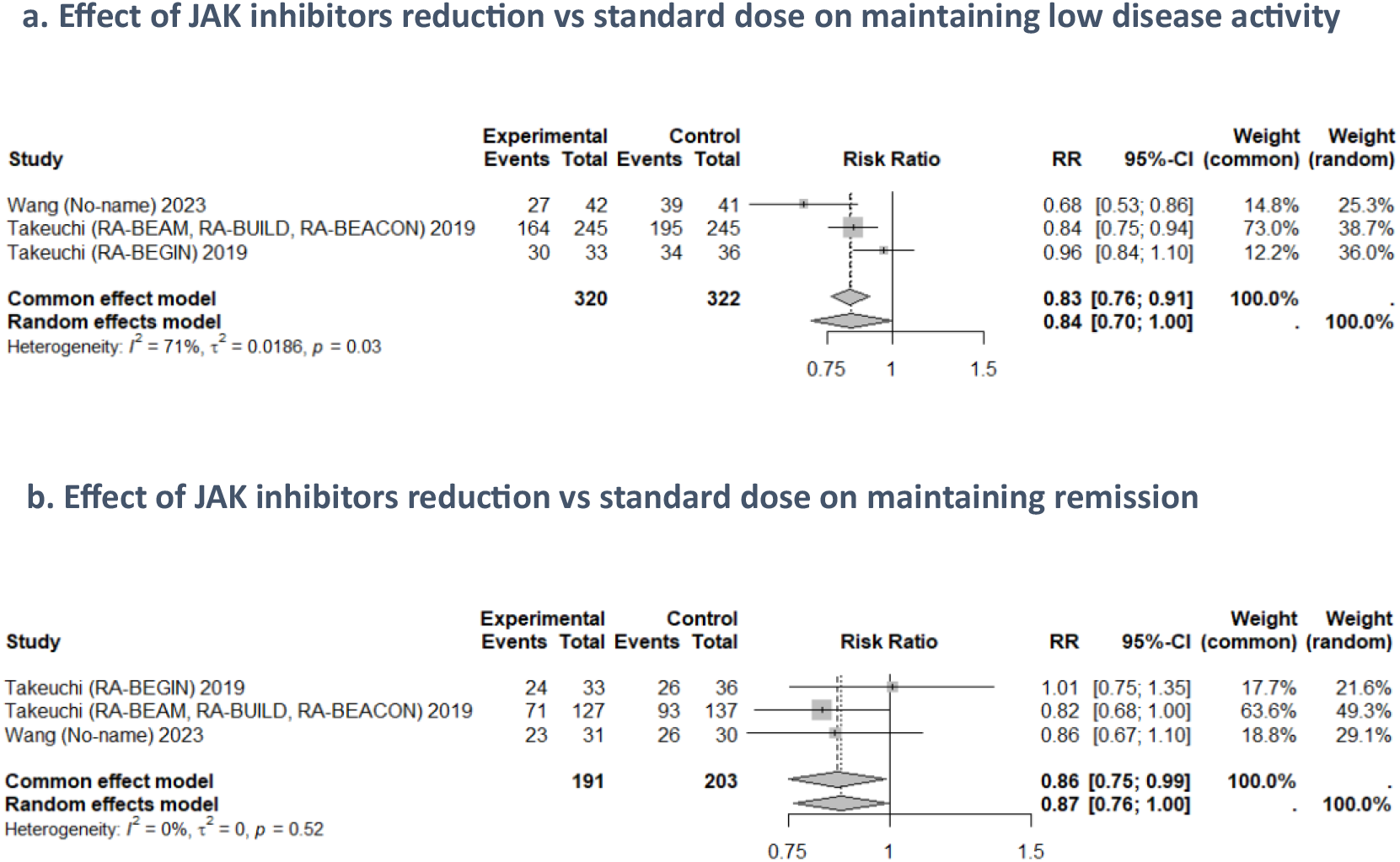
Effect on disease activity comparing JAK inhibitors tapering strategy to maintenance full-treatment. RR: Risk ratio; CI: Confidence interval.

### Outcomes of tocilizumab

A meta-analysis was not performed because the data were not included in the paper.

1. **Remission:** Sanmarti et al. [26] observed more patients on full-dose tocilizumab (90%) remained in remission compared to the RG (73%), p=0.004, based on DAS28-ESR<2.6. Nevertheless, no statistically significant differences were identified when employing alternative efficacy criteria, although a greater number of patients in the SG maintained remission or LDA (Table 1).
2. **Flares-up:** the ToLEDo study [12] reported that 60 patients in the RG experienced flare-ups (42 patients treated with tocilizumab) with a cumulative incidence of 66.6%. In comparison, 26 patients in the SG, with a cumulative incidence of 23.6%. Non-inferiority (NI) was not demonstrated between SG and RG in the overall population or subgroups. In the tocilizumab subgroup, the difference in flare incidence was +41.8% (95% CI 27.0-56.6). More frequent major flares were observed in the RG (15 patients, 8 treated with tocilizumab; cumulative incidence 16.85%) compared to SG (8 patients; cumulative incidence 7.27%). NI for major flares could not be demonstrated in the overall population or the subgroups. For tocilizumab, the difference was +6.7% (95% CI −3.0-16.4).

## Discussion

This manuscript presents an updated systematic review and meta-analysis that includes new RCTs and medications, such as JAK inhibitors and tocilizumab, in addition to anti-TNF therapies. We summarize the evidence from 15 RCTs examining the reduction of biologics and JAK inhibitors compared to the maintenance of standard treatment in adult patients with RA. In comparison with prior systematic reviews, our analysis incorporates seven studies that were conducted more recently (Ruwaard et al. [20], TapERA [22], ARTIC-REWIND [13]), and involving drugs such as tocilizumab (Sanmarti et al. [26], ToLEDo [12]) and JAK inhibitors (Takeuchi et al. [27], Wang et al. [28]), which were not included in earlier reviews. These data suggest that tapering strategies are associated with a disease worsening, defined by a higher risk of flare-ups and a lower likelihood of maintaining remission.

The topic has been previously covered in several systematic reviews. The Cochrane systematic review [7] included 14 studies, eight of which we also reviewed [4,17-19,21,23-25]. All these studies only involved anti-TNF drugs. This review concluded that dose reduction of anti-TNF drugs does not significantly differ from the standard dose in terms of mean DAS28, or persistent remission. However, our results show a lower probability of maintaining remission and a higher risk of flares. This discrepancy in remission could be explained by the fact that the Cochrane review defined persistent remission as patients with LDA. Our data show that tapering does not differ from the standard dose when LDA is analyzed, but discrepancy persists whether remission is the outcome analyzed. Another explanation is that our review, in the maintenance of the remission analysis, includes two RCTs not included in the Cochrane review for this outcome. In concordance with our review, Henaux et al. [29] showed that tapering bDMARDs resulted in a higher risk of losing remission compared with the full dose.

The systematic reviews by Vasconcelos et al. [30] and Henaux et al. [29] showed, as in our study, that there was no significant difference in LDA between maintenance treatment and reduced doses of bDMARDs.

Regarding the tapering of JAK inhibitors (tofacitinib and baricitinib), our systematic review and meta-analysis show that tapering increases the risk of flares and reduces the probability of remission and LDA. However, Wang et al. [28] and Takeuchi et al. [27] noted that tapering results are worse when outcomes are assessed in patients with LDA at baseline compared to those in remission.

In our study, we did not perform a meta-analysis on tocilizumab because the data were not available. The systematic review included two RCTs, Sanmarti et al. [26] and Kedra et al. [12], which showed that patients receiving standard doses of tocilizumab had better results compared to those who tapered.

This systematic review had several limitations. Firstly, the included studies were heterogeneous, with varying primary and secondary outcomes. Secondly, patients were not comparable due to differences in previous treatment duration and disease status across trials. Thirdly, some studies had a relatively short follow-up period, which could limit the reliability of the findings. Additional limitations included diverse study designs, different indices to measure efficacy outcomes, and no direct comparison between SG and RG in studies comprising more than two arms.

Regarding the risk of bias, only four studies were blinded, and two used a PROBE design. In the remaining studies, a higher risk of bias was present; however, this was confined to the outcome measurement domain due to the open-label design and did not impact the overall quality of the studies.

In conclusion, considering that tapering anti-TNF increases the risk of flares and decreases the number of patients in persistent remission, although no difference was observed in maintaining LDA, routine tapering of anti-TNF doses for all RA patients must not be recommended. Similarly, these data do not advocate for routine dose tapering of JAK inhibitors as it leads to fewer patients in persistent LDA and remission, and more frequent and earlier flare-ups. Regarding tocilizumab, the available data also does not suggest routine dose tapering, as it lowers the likelihood of maintaining remission or LDA, and patients experience more relapses.

However, some patients remain in remission or LDA when they reduce their treatment doses, which shows how important it is to identify patients likely to benefit from tapering. This can be done using predictive models based on clinical parameters to anticipate flare-ups or remission [31], however these models have limited efficacy. New predictive models must integrate molecular data (genetic, proteomic, and other omics variables) with clinical data to improve personalized medicine [32]. More controlled studies are necessary, ideally double-blind and NI studies with longer follow-ups, and involving non-anti-TNF.

## Data Availability

All data produced are available online on PROSPERO (CRD42024509208).

https://www.crd.york.ac.uk/PROSPERO/view/CRD42024509208

## Additional Information

Sandra Rotea-Salvo, Laura Galindo-Domínguez, Belén Acasuso Pardo-de-Vera, Vanesa Balboa-Barreiro, Luis Ramudo-Cela, Maite Silva-Díaz, Natividad Oreiro-Villar, Francisco J. De-Toro-Santos, and Isabel Martín-Herranz declare no competing interests. Francisco J. Blanco-García received grant/research support from Sanofi-Aventis, Lilly, BMS, Amgen, Pfizer, AbbVie, TRB Chemedica International, GlaxoSmithKline, Archigen Biotech Limited, Novartis, Nichi-iko pharmaceutical Co, Genentech, Janssen Research & Development, UCB Biopharma, Centrexion Therapeutics, Celgene, Roche, Regeneron Pharmaceuticals Inc, Biohope, Corbus Pharmaceutical, Tedec Meiji Pharma, Kiniksa Pharmaceuticals, Ltd, Gilead Sciences, Inc, consulting fees from Lilly, BMS, Grunenthal, and Pfizer.

## Acknowledgment section

This work has been funded partially by Instituto de Salud Carlos III (ISCIII) by the grant RICORSREI-RD21/0002/0009, European Union-NextGeneration EUPlan de Recuperación transformación y resiliencia and by grant IN607A2021/07 from Xunta de Galicia. SR was supported by ISCIII Programa Rio Hortega CM22/00210 and AG was supported by Catedra FSR-UDC.

## Declaration of generative AI and AI-assisted technologies in the writing process

During the preparation of this work, the authors used ChatGPT and Grammarly to improve readability and language. After using these tools, the authors reviewed and edited the content as needed and took full responsibility for the content of the publication.

## Data availability

The datasets used in this article can be found in the full-text articles included in the systematic review and meta-analysis. The data underlying this article are available in the article and in its online supplementary material.

## Authors’ contributions statement

Sandra Rotea-Salvo: literature search, data collection, data interpretation, writing original draft. Laura Galindo-Domínguez: data collection, data analysis, data interpretation, writing original draft. Belén Acasuso Pardo-de-Vera: data collection, data interpretation, writing review. Vanesa Balboa-Barreiro: data analysis, methodology, writing review. Luis Ramudo-Cela: data analysis, supervision, writing review. Maite Silva-Díaz: data interpretation, writing review. Natividad Oreiro-Villar: data interpretation, writing review. Francisco J. De-Toro-Santos: data interpretation, writing review. Isabel Martín-Herranz: conceptualization, supervision, writing review. Francisco J. Blanco-García: conceptualization, study design, methodology, supervision, writing original draft and review.

