## Supplemental material for "Efficacy of Tapering Biologics and JAK Inhibitors in Rheumatoid Arthritis: A Systematic Review and Meta-analysis"

**Supplementary Table S1: Search strategies of each database.**

| Database | Search strategy |
|---|---|
| Pubmed | ((rheumatoid arthritis[Title/Abstract]) AND (Down-titration[Title/Abstract] OR titration[Title/Abstract] OR taper*[Title/Abstract] OR reduction[Title/Abstract] OR spac*[Title/Abstract] OR de-escalat*[Title/Abstract])) AND (Infliximab[Title/Abstract] OR adalimumab[Title/Abstract] OR etanercept[Title/Abstract] OR certolizumab[Title/Abstract] OR golimumab[Title/Abstract] OR tocilizumab[Title/Abstract] OR tofacitinib[Title/Abstract] OR baricitinib[Title/Abstract] OR upadacitinib[Title/Abstract] OR filgotinib[Title/Abstract] OR janus kinase inhibitor[Title/Abstract] OR jak inhibitor*[Title/Abstract] OR "Tumor Necrosis Factor Blockers"[Title/Abstract] OR tnf inhibitors[Title/Abstract] OR "Tumor Necrosis Factor Antagonist"[Title/Abstract] OR "Tumor Necrosis Factor Inhibitor"[Title/Abstract] OR tnf antagonist[Title/Abstract]) Filters: Clinical Trial, Randomized Controlled Trial |
| Embase | ("rheumatoid arthritis" and (Infliximab or adalimumab or etanercept or certolizumab or golimumab or tocilizumab or tofacitinib or baricitinib or upadacitinib or filgotinib or "janus kinase inhibitor" or "jak inhibitor*" or "Tumor Necrosis Factor Blockers" or tnf inhibitors or "Tumor Necrosis Factor Antagonist" or "Tumor Necrosis Factor Inhibitor" or "tnf antagonist") and (Down-titration or titration or taper* or reduction or spac* or de-escalat*)).mp. [mp=title, abstract, heading word, drug trade name, original title, device manufacturer, drug manufacturer, device trade name, keyword heading word, floating subheading word, candidate term word]. Limit to (human and (clinical trial or randomized controlled trial)) |
| Web of Sciences | ((TS=(rheumatoid arthritis)) AND TS=(Infliximab OR adalimumab OR etanercept OR certolizumab OR golimumab OR tocilizumab OR tofacitinib OR baricitinib OR upadacitinib OR filgotinib OR janus kinase inhibitor OR jak inhibitor* OR "Tumor Necrosis Factor Blockers" OR tnf inhibitors OR "Tumor Necrosis Factor Antagonist" OR "Tumor Necrosis Factor Inhibitor" OR tnf antagonist)) AND TS=(Down-titration OR titration OR taper* OR reduction OR spac* OR de-escalat*) and Clinical Trial (Document Types) |
| Cochrane Library | (rheumatoid arthritis):ti,ab,kw AND (Down-titration OR Titration OR Taper* OR de-escalate* OR reduction OR spac*):ti,ab,kw AND (infliximab OR adalimumab OR etanercept OR certolizumab OR golimumab OR tocilizumab OR tofacitinib OR baricitinib OR upadacitinib OR filgotinib OR janus kinase inhibitor OR jak inhibitor* OR "Tumor Necrosis Factor Blockers" OR tnf inhibitors OR "Tumor Necrosis Factor Antagonist" OR "Tumor Necrosis Factor Inhibitor" OR tnf antagonist):ti,ab,kw en Ensayos (Se han buscado variaciones de la palabra) |
